# Durability of pulmonary vein isolation and feasibility and safety of additional intrapulmonary vein sleeve ablation by means of a circular pulsed-field ablation catheter. The PVI-PVA study

**DOI:** 10.1101/2025.10.22.25338592

**Authors:** Dimitrios Tsiachris, Athanasios Kordalis, Christos-Konstantinos Antoniou, Aikaterini-Eleftheria Karanikola, Ioannis Doundoulakis, Aggeliki Laina, Panagiotis Xydis, Konstantinos Tsioufis

**Author notes:** **Corresponding author:** Dimitris Tsiachris Vasilissis Sofias 114, ave Athens, Greece, 11527.

## Abstract

**Background:** Pulmonary vein isolation (PVI) constitutes the established strategy for atrial fibrillation (AF) ablation. With the advent of PV stenosis risk-free pulsed field ablation (PFA), we explored the feasibility and safety of additional ablation within PV sleeves (PVA). Moreover, we assessed the durability of PVI through PFA in a deep sedation setting, comparing a 3D Electroanatomical Mapping (3D-EAM)-based navigation approach with the standard fluoroscopy-based one.

**Methods:** In this single-center, first-in-human study (NCT07035288), 40 AF patients underwent first time PFA-based PVI+PVA (4 additional applications inside the PVs) between November 2024 and April 2025 using a circular array PFA catheter (PulseSelect, Medtronic, Minneapolis, MN), randomized to either 3D-EAM-based (Carto3 prime, J&J, Irvine, CA) or fluoroscopy-based navigation, in a propofol-based deep sedation setting. Rate of conversion from 3D-EAM to fluoroscopy-based navigation was recorded. First-pass isolation was assessed immediately post-ablation. Venography and 3D-EAM were performed at remapping at 2 to 3 months after the index procedure.

**Results:** No major procedure-related adverse events were noted, as well as no acute kidney injury, significant hemolysis or phrenic nerve palsy. First-pass isolation was successfully achieved in 95% of patients (3D-EAM: 95%, fluoroscopy: 95%, p=NS) and in 98.7% of PVs. Venography performed at remapping revealed no PV stenosis. PVI durability per patient was 92.5% (3D-EAM: 90%, fluoroscopy: 95%, p=NS) and per vein was 97.5% (3D-EAM: 97.5%, fluoroscopy: 97.5%, p=NS between navigation methods). Half of 3D-EAM cases were converted to fluoroscopy due to map shift.

**Conclusions:** PFA-based PVA is safe. Catheter performance, as depicted by 97.5% durable PVI, was such that adding a 3D-EAM system was not associated with improved efficacy and exhibited high conversion rate to fluoroscopy-based navigation in a deep sedation setting.

**Categories:** electrophysiology

**Special Sections:** Original article

**Randomized clinical trial:** NCT07035288

## Introduction

Atrial fibrillation (AF) is the most common cardiac arrhythmia in clinical practice^1^. It is well recognized that AF can cause stroke, congestive heart failure and systemic thromboembolism, all of which result in high mortality and poor quality of life^1^. Catheter ablation is superior to drug therapy in maintaining sinus rhythm^1, 2^ and is associated with improved ejection fraction, NYHA status and survival, compared with medical therapy in patients with AF and cardiovascular risk factors^3–5^.

Electrical isolation of the pulmonary veins (PVs) is the base of AF ablation and is currently achieved using a technique referred to as PV isolation (PVI)^1, 2^. Thermal radiofrequency (RF) ablation within the PVs was initially used to target PV triggers themselves, though accompanied by poor efficacy and PV stenosis^6^. Subsequently, segmental ostial ablation was adopted, limiting ablation deep within the PVs without avoiding PV stenoses^7, 8^. Ultimately, antral PV isolation was finally designated and is currently widely used as it allowed elimination of PV stenosis along with reduced AF recurrence^9^. Antral PVI has since become the standard both for point-by-point thermal ablation and one-shot technologies^1, 2^.

However, the rationale for targeting the potentials of the intrapulmonary vein myocardial sleeves remains valid^10^. Pulmonary vein reconnection remains the most important cause of AF recurrence after initial AF ablation^1, 2^; thus, achieving better durability is a *sine qua non* for improving patient outcomes. Pulsed field ablation (PFA) is a novel energy form, characterized by relative myocardial selectivity at low voltages, and, most relevant to our endeavor, lower risk for pulmonary vein stenosis^11–13^, a serious iatrogenic complication with significant morbidity^14^. Within a few years, PFA has achieved noninferiority^15, 16^, and starts to exhibit, in certain populations, superiority to thermal-based ablation^17^. The PulseSelect catheter (Medtronic, Minneapolis, MN) is an approved PFA technology for AF ablation^15^. PVI by means of the PulseSelect catheter is performed under either general anesthesia or deep sedation^18^ and can be guided by fluoroscopy^19, 20^ or, alternatively, the catheter can be visualized through 3-dimensional electroanatomical mapping (3D-EAM), facilitating accuracy of lesions^19, 20^. Consequently, with the advent of PV stenosis risk-free PFA, we aimed to explore the feasibility and safety of additional ablation targeting electrically active PV sleeves (pulmonary vein ablation – PVA) by means of the PulseSelect catheter^13^. Moreover, we assessed durability of PVI through PFA in a deep sedation setting comparing a 3D-EAM-based navigation approach with the standard fluoroscopy-based navigation approach acutely and long term (remapping at 2-3 months).

## Methods

### Study design

In this single-center, first in human study (NCT07035288), 40 AF patients were submitted to a PFA-based PVI+PVA approach between November 2024 and April 2025 using a circular array PFA catheter (PulseSelect, Medtronic, Minneapolis, MN), randomized one to one to either 3D-EAM-based (Carto3 prime, J&J, Irvine, CA) or fluoroscopy-based navigation, in a propofol-based deep sedation setting, with blinded adjudication of endpoints.

### Study population

Inclusion criteria – All of the following criteria had to be fulfilled:

1. Patients aged ≥18 and <80 years old.
2. Patients with symptomatic paroxysmal or persistent AF requiring PVI.
3. Signed written informed consent by the patient for participation in the study and agreement to comply with the procedure and the follow-up schedule

Exclusion Criteria – Presence of one of:

1. Patients with previous AF ablation
2. Ejection fraction<35%, coronary artery bypass surgery within the previous 3 months, transient ischemic attack or stroke within the previous 6 months
3. Any contraindication to oral anticoagulation.
4. Any contraindication to deep sedation.

Participants first received oral anticoagulation for 3 weeks. Thereafter patients were randomized either to fluoroscopy-based or 3D-EAM-based PVI+PVA by means of the PulseSelect catheter. Oral anticoagulation was continued until the morning of the procedure and resumed on the evening after the intervention.

### Sedation protocol

All ablation procedures were performed using the same deep sedation using boluses of midazolam, fentanyl and propofol administered by a nurse under the supervision of the electrophysiologist. Hemodynamic and respiratory monitoring was performed by the personnel of the electrophysiology laboratory staff monitoring continuous heart rate, invasive blood pressure, peripheral capillary oxygen saturation and respiratory rate. All staff members were trained in airway management. Exclusion criteria for the use of this protocol were a body mass index >35 kg/m2 and severe untreated sleep apnea syndrome.

Sedation was initiated with an intravenous bolus injection of 1-2mg of midazolam plus 20-30mg of propofol with a maximum of 50mg. After 10 minutes of initial assessment of sedation, additional boluses of 10-20mg of propofol (maximum of 25mg) were administered in order to maintain deep sedation by the operator through the side port of the sheath. Prior to the PFA energy applications, the operator administered an intravenous bolus of 0.5mg atropine as well as of lidocaine of 1 mg/kg to suppress a coughing response.

### Ablation protocol

Twenty patients were randomized to 3D-EAM-based and 20 to fluoroscopy-based navigation. Baseline venography was first performed in all cases by using the steerable sheath (FlexCath Contour) for contrast infusion into each PV antrum. The images were stored to improve PFA catheter positioning in the fluoroscopy-guided group and served as baseline images.

#### a. Fluoroscopy-guided pulmonary vein isolation and ablation (PVI+PVA)

The PulseSelect circular electrode array catheter has a 20° forward tilt with electrodes 4-5-6 most distally located, which may contact the tissue first, assuming a toroidal configuration (at least upon conventional use). Thus, the catheter delivers a horseshoe shaped lesion. Circumferential vein ablation is achieved by rotating the catheter between ablations and flexing the sheath to create overlapping lesion segments. Electrode 5, always distally located, is used as a reference during successive 45° rotations between lesions. Electrode spacing is fixed and produces a reliable field for predictable and consistent contiguous energy delivery. One application (4 pulse trains) was delivered at each catheter position, and a minimum of 8 applications were completed for each vein, ensuring continuous and overlapping lesions (4 ostial and 4 antral). Concerning PVA, an additional set of 4 lesions inside the pulmonary veins were performed *before* the ostial and antral lesions. The PulseSelect catheter was partially deployed, allowed to assume a helical configuration adjusted to PV shape and ensuring contact with their wall. Subsequently, the catheter was rotated within each PV, moving from deeper to shallower parts, in order to avoid overlap (Figure 1).

**Figure 1.**
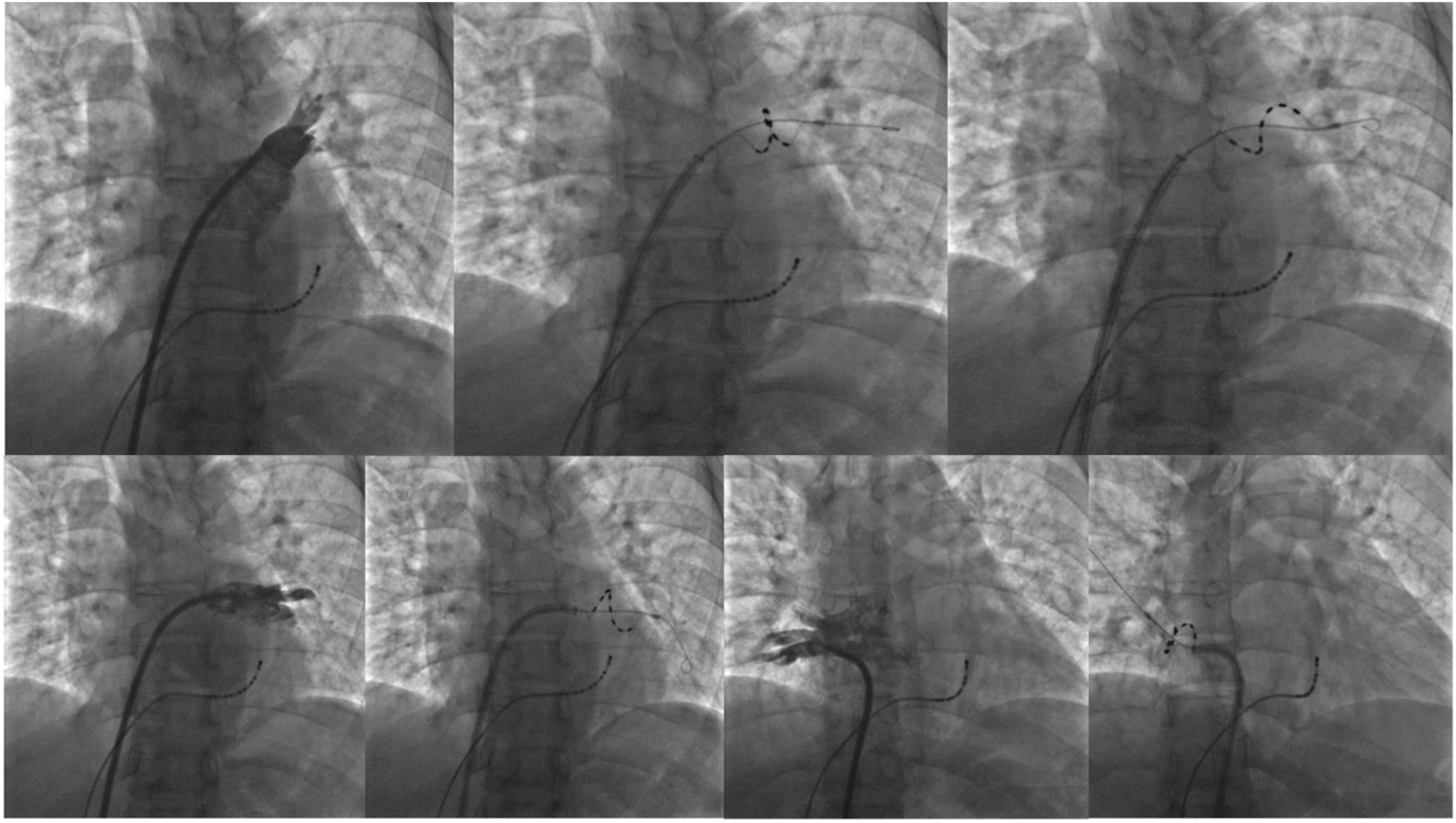
Clockwise from top left: Venogram of left superior pulmonary vein and subsequently fluoroscopic images of catheter conformation for delivery of PVA lesions. PVA lesion in the right inferior pulmonary vein, next to its venogram, and, finally, PVA lesion in the left inferior pulmonary vein and its corresponding venogram. The helical configuration and careful coaxial advancement are evident. Finally, in the left superior vein the rotation between the delivery of the 2 lesions sets is evident (PVA – pulmonary vein ablation).

#### b. PVI combined with electroanatomical mapping (EAM)

The Carto 3 Prime system (J&J, Irvine, CA) was used for visualization of the PulseSelect catheter. A multipolar catheter (PentaRay, J&J, Irvine, CA) was used to map the left atrium pre-ablation in the 3D-EAM-based navigation group. Flexion and rotation of the sheath (FlexCath Contour®) is used for initial positioning and rotation of the catheter shaft for pivoting the loop and ablating in successive steps in a rotational pattern. During catheter manipulation, the full array is visualized, and the fifth electrode is highlighted and used as quick real-time reference. Snapshots of the array immediately prior to pulse delivery are used as reference for planning subsequent ablation sites. In cases of an extreme angle of the right inferior pulmonary vein, placement of the PulseSelect catheter in the bottom part of the vein without the guidewire inside the vein may be attempted by means of 3D-EAM and monitoring the fifth electrode of the catheter. Need for conversion from 3D-EAM-based to fluoroscopy-based navigation due to map shifting was recorded.

### Immediate remapping

First-pass isolation was assessed by means of Carto 3 Prime® 3D-EAM immediately post-ablation in both navigation method groups. All veins were reassessed for bidirectional block to confirm continued electrical isolation. Any gaps identified were targeted and ablated with an RF catheter (Navistar STSF J&J Irvine CA). The extent of isolation in terms of surface area was measured, as well as any extension of isolation in sites of anatomical interest (posterior wall, roof, carinas, anterior wall, ridge between left PVs and left atrium appendage). Specifically, we assessed left and right antral perimeter, and the antral to antral distance in the roof, posterior part and floor of the antra.

### Follow up study

All patients were re-evaluated 2-3 months post-ablation. Changes of therapy were not allowed during the blinding period of 2 months. Clinical atrial tachyarrhythmia recurrence, need for electrical cardioversion and need for hospitalization were recorded. In all patients, the left atrium was accessed, and contrast was injected within each PV through the same sheath type as in the index procedure (FlexCath Contour®). Resulting images were then compared with baseline images before PVI+PVA.

Post-ablation remapping was carried out in the same way as the 3D-EAM in the index procedure. Again, all veins were reassessed for bidirectional block to confirm durable electrical isolation. To assess PV reconnection and gap localization, a high-density voltage map was acquired with the color display range set initially to 0.20-0.50mV, then reduced to 0.10-0.40mV to accentuate the border zone between healthy tissue and scar, facilitating visual identification. Gaps were classified according to each PV quarter (anterior, superior, posterior, inferior). Stored fluoroscopy and electroanatomical mapping images from the primary procedure were used for analysis. In case of PV reconnection, the earliest activation site upon pacing from the nearby left atrium was identified as a potential gap location. Any gaps identified were targeted and ablated with RF. Finally, we assessed durability of ablation surface measuring left and right antral perimeter, and antral to antral distances in the roof, posterior part and floor of the antra in comparison to respective initial measurements obtained immediately post ablation.

### Endpoints and objectives

Primary and secondary endpoints were used to assess the comparative data of methodological alternatives as then related to safety, efficacy, and efficiency outcomes, and they include.

Primary Objective(s) / Endpoint(s):

- Safety of PFA lesions inside the PVs, both acutely and upon remapping 2-3 months post-ablation (assessment for PV stenosis).
- Durability of PVI based on remap (percentage of patients with all PVs isolated, percentage of PVs isolated).
- Comparison of PVI durability between the 2 navigation approaches at index procedure (fluoroscopy-based and 3D-EAM based).
- Major acute complications of ablation (death, cardiac tamponade not related to transseptal puncture). Time frame: Acute peri-procedural complications were defined as occurring within 24 hours of ablation.

### Secondary Objective(s) / Endpoint(s)

- Identification of PVI gaps acutely based on immediate 3D-EAM remapping (first-pass isolation).
- Identification of PVI gaps on remapping 2-3 months post-index procedure.
- Rate of conversion from 3D-EAM to fluoroscopy-based navigation due to map shifting.
- Fluoroscopy time and dose, procedure time, time from left atrium access time to sheath removal, left atrium dwelling time.
- Major acute complications of deep sedation and propofol use (anesthesiologist called, intubation, admission to ICU).
- Minor complications of ablation (acute kidney injury, significant hemolysis or phrenic nerve palsy)

### Data and Statistical Plan

All stats were conducted using R software. All safety and efficacy outcomes were summarized using descriptive statistics. Parametric variables were assessed for normality of distribution using the Shapiro-Wilk test. All normally distributed parametric variables were described using the mean ± standard deviation approach. All non-normally distributed parametric variables were described using the median (25^th^-75^th^ percentile) expression. Categorical values were presented as frequencies and percentages.

Student’s *t*-test was used for assessing differences between 2 normally distributed parametric variables, while the Mann-Whitney U-test was used for comparing differences between 2 non-normally distributed parametric variables. Chi-square test was used for comparisons between categorical variables, with Fisher’s exact test substituted in cases with expected cell frequencies<5. All procedural parameters and analyses are made on an “intention-to-treat” basis. A p-value <0.05 was considered to indicate statistically significant differences in all cases. No statistical adjustment was made for the multiple secondary endpoints in the analysis.

## Results

Forty patients with AF [age: 65.5 (58-71) years, 74% males, 19 (48%) paroxysmal, 21 (52%) persistent] underwent treatment under deep sedation with the circular PFA catheter [3D-EAM-based: 20 (50%)]. Mean CHA_2_DS_2_-VASc score was 1.6±1.3 and no patient has experienced a cerebrovascular event (stroke / transient ischemic attack). Mean ejection fraction was 55.5 ±7.7%. Mean left atrial diameter was 43.7±1 mm. Six patients (15%) had known obstructive sleep apnea. Median body mass index was 27.6 (24.9-29.7) kg/m^2^. Sixty percent of patients were on antiarrhythmic medications (either class Ic or class III) (Table 1). Map shifting due to patient movement was noted in half of the EAM-based group during PFA applications, resulting in continuation of the ablation based on fluoroscopy and baseline venography images. There were no statistically significant differences between navigation method-based groups regarding age, sex, CHA2DS2-VASc score, ejection fraction, left atrial diameter, and vascular disease, hypertension and type II diabetes rates (Table 1).

**Table 1.** Baseline demographic data.

| Variable | All (n=40) | 3D-EAM (n=20) | Fluoroscopy (n=20) | p-value |
| --- | --- | --- | --- | --- |
| Age (years) | 65.5(58-71) | 65.5(57.5-71) | 65.5(60-72) | 0.63 |
| Sex (% males) | 75% | 70% | 80% | 0.72 |
| BMI (kg/m<sup>2</sup>) | 27.6 (24.9-29.7) | 27.4 (24.7-31.5) | 27.8 (24.9-29.4) | 0.98 |
| CHA2DS2-VASc score | 1.63±0.2 | 1.7±0.27 | 1.55±0.3 | 0.71 |
| Hypertension (% yes) | 50% | 50% | 50% | 1 |
| Diabetes type 2 (% yes) | 22.5% | 25% | 20% | 1 |
| Coronary/Peripheral artery disease (% yes) | 10% | 10% | 10% | 1 |
| Stroke/TIA (% yes) | 0% | - | - | - |
| Obstructive sleep apnea (% yes) | 15% | 20% | 10% | 0.66 |
| Atrial fibrillation type (% paroxysmal) | 47.5% | 55% | 40% | 0.53 |
| Anticoagulation (% yes) | 97.5% | 100% | 95% | 1 |
| Antiarrhythmics (% yes) | 60% | 50% | 70% | 0.73 |
| Ejection fraction (%) | 55.5±7.7 | 54.3±8.9 | 56.7±6.3 | 0.57 |
| Left atrial diameter (mm) | 43.7±1 | 43.8±2 | 43.6±0.9 | 0.92 |

Regarding procedural times, average procedure duration was 80±2.4 minutes. Due to a high conversion rate from 3D-EAM-guided to fluoroscopy-guided ablation (10 patients – 50%), concerning procedural times, along the standard “intention to treat”, an “as treated” analysis was performed as well (Table 2). More specifically, in patients with “true” 3D-EAM-guided ablation mean procedural duration was 76±10 minutes as opposed to 82±15 minutes in the fluoroscopy (per intention or by conversion)-guided group (p=0.29, no statistically significant difference). Left atrial catheter dwelling time was 39±6.7 minutes in the “true” 3D-EAM-guided group, as opposed to 49±14 minutes in the fluoroscopy-guided one (p=0.04 – statistically significant, favoring the 3D-EAM-guided approach). Finally, regarding fluoroscopy time (as treated analysis), the relevant values were 8.8±5.3 minutes (“true” 3D-EAM) and 12.5±5.2 minutes (fluoroscopy), yielding a p-value of 0.06, favoring 3D-EAM and narrowly missing statistical significance. Regarding total post ablation mapping time, it averaged 12.2±0.6 minutes (p=0.69 between intention to treat groups) – an “as treated” analysis was not carried out because all maps were constructed de novo, i.e. without keeping the initial anatomical shell in the 3D-EAM-guided approach.

**Table 2.** Procedural times by navigation method-based groups.

| Intention to treat analysis |  |  |  |  |
| --- | --- | --- | --- | --- |
| Variable | All (n=40) | 3D-EAM (n=20) | Fluoroscopy (n=20) | p-value |
| Procedure duration (min) | 80±2.4 | 82±3.2 | 79±3.7 | 0.52 |
| Left atrial dwelling time (min) | 46.5±2.3 | 47.9±3.6 | 45.4±3.1 | 0.6 |
| Total fluoroscopy time (min) | 11.6±3.9 | 10.94±4.52 | 12.18± 3.25 | 0.76 |
| Post-ablation mapping time (min) | 12.2±0.6 | 12.5±1.24 | 11.9±0.78 | 0.69 |
| As treated analysis |  |  |  |  |
| Variable | "True" 3D-EAM (n=10) | Fluoroscopy / conversion (n=30) | p-value |  |
| Procedure duration (min) | 76±10 | 82±15 | 0.29 |  |
| Left atrial dwelling time (min) | 39±6.7 | 49±14 | 0.04 |  |
| Total fluoroscopy time (min) | 8.8±5.3 | 12.5±5.2 | 0.06 |  |

### Safety data

Regarding PVI+PVA acute safety (Table 3), no major or minor procedure-related adverse events were noted, including PV stenosis, tamponade, stroke, phrenic nerve injury, esophageal damage, acute kidney injury, clinically significant hemolysis or mortality. Importantly, no catheter entrapment was observed during PVA through the helical position of the circular catheter. Venography performed during remapping at 2-3 months revealed no PV stenosis. First-pass isolation was successfully achieved in 95% of patients (3D-EAM: 95%, fluoroscopy: 95%, p=NS) and in 157/159 (98.7%) of pulmonary veins (3D-EAM: 98.7%, fluoroscopy: 98.75%, p=NS between navigation methods – Table 3).

**Table 3.** Procedural outcomes at remapping.

| Variable | All (n=40) | 3D-EAM<br>(n=20) | Fluoroscopy<br>(n=20) | p-value |
| --- | --- | --- | --- | --- |
| Pulmonary vein stenosis (number of veins) | 0 | - | - | - |
| Mortality (n dead) | 0 | - | - | - |
| Any one of: tamponade, stroke, phrenic nerve palsy, esophageal damage, acute kidney injury, significant hemolysis (n) | 0 | - | - | - |
| First-pass isolation per patient (n successful) | 95% (38/40) | 95% (19/20) | 95% (19/20) | 1 |
| First pass isolation per vein (n successful) | 98.7% (157/159) | 98.7% (78/79) | 98.75% (79/80) | 0.95 |
| Pulmonary vein isolation durability at remapping, per patient (n successful) | 37 | 90% (18/20) | 95% (19/20) | 1 |
| Pulmonary vein isolation durability at remapping, per vein (n successful) | 97.5% (155/159) | 97.5% (77/79) | 97.5 (78/80) | 1 |

### Durability data

At 2 to 3 months, PVI durability per patient was 92.5% (3D-EAM: 90%, fluoroscopy: 95%, p=NS) and per vein was (155/159) 97.5% (3D-EAM: 97%, fluoroscopy: 97.5%, p=NS between navigation methods) (Table 3). Of the 4 reconnected veins requiring touch-up RFA, a single vein was reconnected in 2 patients, and 2 veins were reconnected in the last patient. The gap was detected in the posterior and inferior part of the right inferior PV in 3 cases and in the inferior part of left inferior PV in the last case. Interestingly, and relevant to the validity of our approach, there were also 4 cases with viable myocardial tissue in the area of wide carinas in the presence of both exit and entrance block in the relative PVs, possibly due to successful PVA (Figure 2).

**Figure 2.**
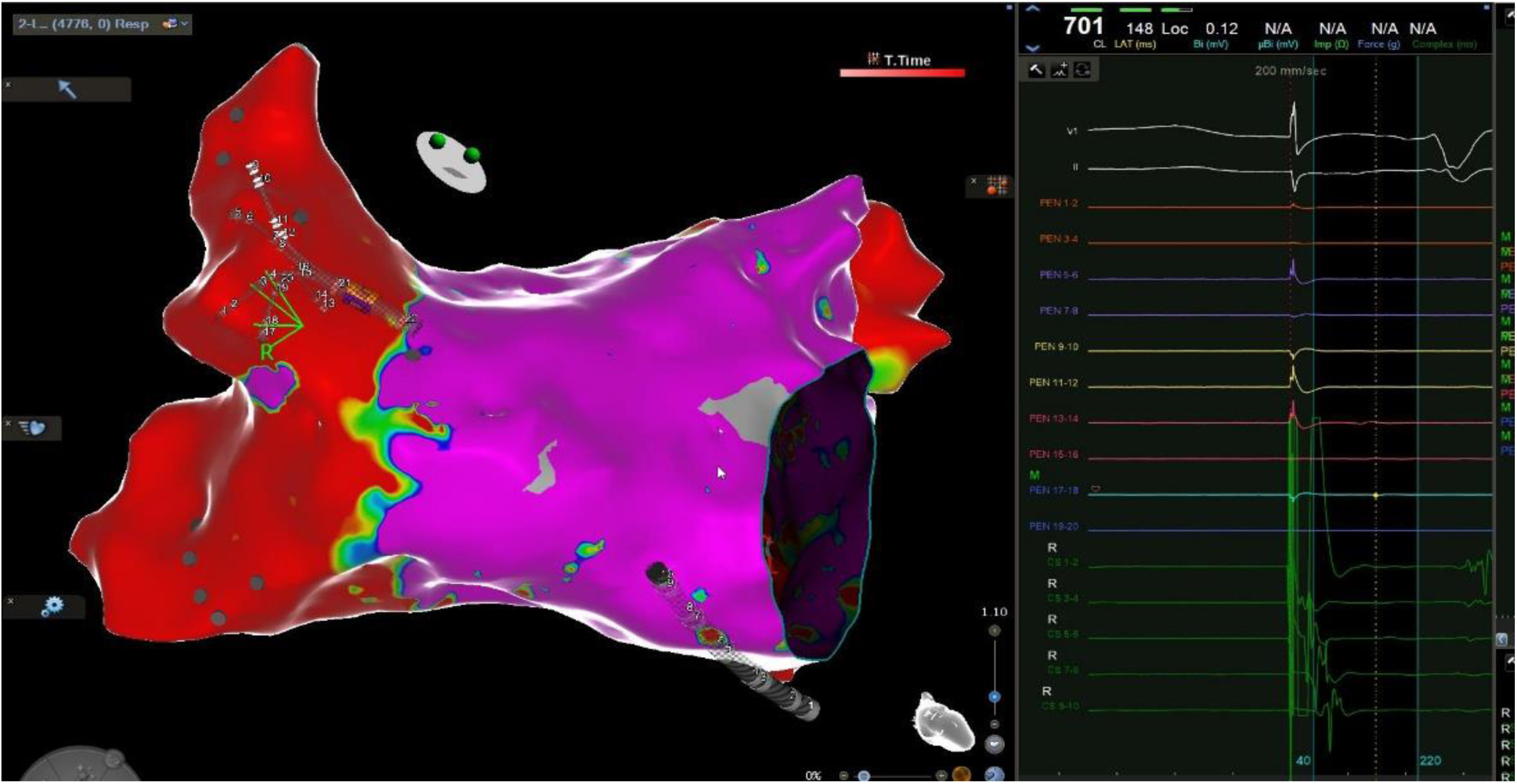
Despite the presence of viable tissue in the right carina entry block in the right superior pulmonary vein is shown on the right. These cases can be considered proof of our PVA incorporation in Af ablation concept.

Focusing on durability of ablation surface we found no significant differences between baseline and follow-up remapping regarding left and right antral perimeters (Table 4). Though the use of analysis of variance (ANOVA), it was shown that there were no statistically significant differences regarding the reduction of ablated surface area between baseline and follow up in terms of navigational method-based groups. In contrast, increases to interantral distances were seen in the roof (by 6 mm, p<0.05) and the inferior part (by 4.6 mm, p<0.05); yet not in the posterior part possibly attributed to better contact catheter in the respective wall. The fact that there were 2 cases in the fluoroscopy group and one case in the EAM group where incidental posterior wall ablation was identified during immediate remapping and persisted during remapping at 3 months is worth mentioning (Figure 3).

**Figure 3.**
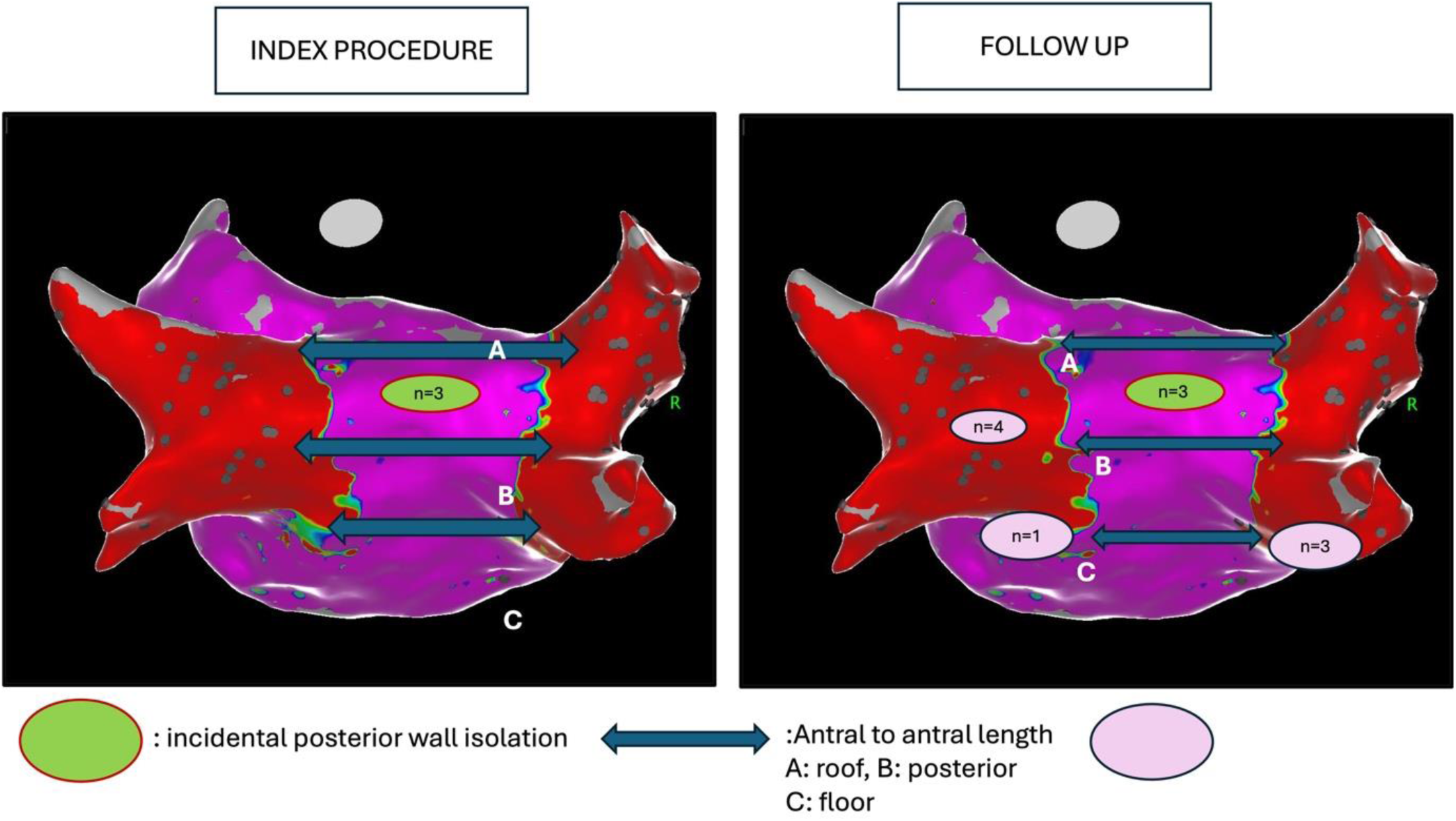
Durability of PVI based on behavior of interantral distance in the posterior wall of the atrium – substrate map in a posteroanterior orientation. Regarding the roof and the inferior distances (lines A and C), significant increases were seen (i.e. the width of the PVI was reduced). In contrast, no significant changes were seen regarding PVI width at the level of line B (posterior part) possibly attribute to better catheter contact. There were 3 incidental posterior wall ablations and, in total, 4 cases with PVA and bidirectional block despite the presence of viable tissue in wide carinas (3 in the right carina and 1 in the left carina).

**Table 4.** Ablation Extension and robustness.

| Variable | Baseline (n=40) | Follow up (n=40) | p-value |
| --- | --- | --- | --- |
| Left Antral Perimeter (mm) | 138.1 (122.7 – 143.9) | 139 (129.1 – 159.2) | 0.08 |
| Right Antral Perimeter (mm) | 142.2 (133.4 – 163.4) | 144.8 (130.9 – 161.7) | 0.98 |
| Antral to Antral distance (mm): |  |  |  |
| @roof | 23 (13.5 – 31.8) | 29 (16.3 – 36.5) | <0.05 |
| @posterior | 24.2 (14.5 – 32) | 28 (12.8 - 38) | 0.21 |
| @floor | 24.3 (15.8 – 29.4) | 28.9 (19 – 36.9) | <0.05 |
| Incidental posterior wall isolation (%) | 3 (7) | 3 (7) | 1 |

## Discussion

In the current work, we presented the use of a circular electrode array PFA catheter for both PV isolation and ablation within the PV sleeves in the context of paroxysmal and persistent AF, in a deep sedation setting. Furthermore, we randomized our cohort into either 3D-EAM or fluoroscopy-based navigation. Our main goal was to assess feasibility, safety, and durability of the PFA PVI+PVA paradigm for AF ablation, with assessment of the benefit, or lack thereof, of using a 3D-EAM system for navigation being a secondary goal.

Initially, one should bear in mind that, as reported in the seminal 1998 paper by Haïssaguerre *et al*^21^ the triggering foci for AF are located at the myocardial sleeves inside the PVs and were themselves targeted in all initial approaches to AF ablation. It was only due to reduced efficacy and PV stenosis occurrence^6^ that the PV isolation approach was pursued instead^9^. However, being able to ablate *inside* the pulmonary veins, targeting the areas of endocardium and endothelium confluence, without concern for causing stenosis, is both a valid approach^10^ and has two-fold effects. To begin with, given the helical configuration of the catheter and its rotation inside the vein, it will lead to an additional isolation level. On the other hand, PVA may prevent arrhythmia recurrence even if PVI is not fully successful, given that the triggering foci themselves have been ablated – proof of concept provided by the 4 patients with viable tissue in their (wide) carinas but with bidirectional PV electrical block, i.e. block without antral PVI. Finally, it could be argued that a future focus on more meticulous PVA and more ostial PVI, instead of wider PVI alone, might reduce the occurrence of atrial tachycardias post-AF ablation (rates ranging from 5%-40%) due to extensive atrial substrate modification, which are often associated with higher morbidity in terms of symptoms and usually require a redo procedure due to poor response to medications^22^.

PFA, as an energy form, is uniquely poised as a PVA strategy due to its graded tissue effect (according to peak voltage)^15–17, 23^. It is not known, though, what effects on electrical field shape the helical compared to toroidal configuration during PVA incurs. Use of the PulseSelect catheter has the advantage that the distance between the electrodes does not change between circular and linear and therefore the field shape should be similar.

Thus, our work could pave the way for a more comprehensive study of PVA by means of PFA and the potential for shifting the paradigm for AF ablation. Indeed, our results suggest at least non-inferiority of the PVI+PVA approach in terms of feasibility, safety, duration, and clinical effectiveness. Unfortunately, including a cohort receiving PVA alone would be extremely difficult due to the unapproved nature of the intervention – although some advantages of PVA may be expressed when PVI is absent (*e.g.* avoiding atrial tachycardias and sparing more atrial myocardial tissue).

Regarding procedural simplification, deep sedation or general anesthesia have been utilized from various centers when using the PulseSelect® system^18^. Each method is associated with distinct advantages. General anesthesia allows for use of paralytics that prevent skeletal muscle spasm that may accompany PFA lesion delivery. Additionally, patient immobilization ensures catheter stability and mitigates the risk of map shifts when EAMs are used. Similarly, paralytics prevent activation of the phrenic nerve during delivery of PFA lesions around the right superior pulmonary vein that can result in a similar effect. Lastly, general anesthesia provides better hemodynamic control during the profound vagal response that often accompanies delivery of PFA lesions to the left-sided veins. In contrast, additional time for induction and need for an anesthesiologist are the main drawbacks of general anesthesia.

If deep sedation is utilized^24^, pretreatment with atropine given intravenously has been proven effective in preventing such vagal response. Anesthesia placement of esophageal temperature probes is not necessary as PFA does not cause significant heating of myocardial or adjacent tissues^25^. Propofol administration is necessary for deep sedation and requires the presence of an anesthesiologist in most (but not all) European centers. Specifically, in many countries such as France, Spain, or Italy, it is not legally possible to perform deep sedation without the presence of an anesthesiologist inside the operating room, whereas in other countries, such as Germany, deep sedation with propofol has been implemented in standard protocols for AF ablation without anesthetists since several years ago. In a worldwide survey^26^ before PFA use, 60% of centers reported that the presence of an anesthesiologist was required to perform deep sedation. Deep sedation, including propofol use without an anesthesiologist might have a role when resources are lacking. Our findings using a deep sedation protocol imply that, although durable PVI and first-pass PVI are not compromised, the potential for patient movement (conscious, unconscious, or iatrogenic) is associated with a 50% rate of conversion from a 3D-EAM to a fluoroscopy-guide navigation – a mapping *and* ablating catheter might have a distinct advantage in this regard, allowing for several “segmental maps” if movement occurred.

Combinational use of a PFA and a 3D-EAM system may increase lesion targeting accuracy allowing for, *e.g.*, ablation of the posterior wall. Additionally, the potential to depict previous lesion areas (by displaying catheter position recorded during previous energy deliveries) facilitates overlapping and reduces the possibility for gaps, which, when present, may be detected during remapping. Indeed, the PulseSelect® catheter is virtually “plug and play” in this regard, given that it is compatible with all available 3D-EAM systems. However, even in cases where conversion to fluoroscopy-based navigation did not occur, 3D-EAM did *not* confer an advantage regarding PVI, first-pass PVI or lesion durability. The only benefit was seen concerning left atrial dwelling and irradiation times. Although counterintuitive, it could be argued that the inclusion of a PVA set of lesions (as discussed above) and the effectiveness of the toroidal catheter configuration, along with ease of shaft manipulation (*i.e.* the PFA system as a whole), were so effective that they rendered moot any potential benefit from 3D-EAM use.

## Study limitations

This is a proof-of concept study enrolling a small number of patients, not designed to test non-inferiority of PVI+PFA. However, as mentioned, it is also a hypothesis-generating study, in the sense of creating interest in the PFA approach. Another limitation lies in the absence of a PVA-only group. We cannot also exclude potential arcing in case of electrodes overlapping. Finally, we cannot generalize our findings to other PFA catheters, given that some one-shot designs do not physically allow for intrapulmonary vein myocardial sleeve ablation. Focal PFA catheters on the other hand may benefit from such an approach as PVA may prove an easier and less time-consuming ablation target.

## Conclusions

Pulsed field ablation using a circular electrode array catheter is safe and effective in achieving durable PVI+PVA for the treatment of AF in a deep sedation context. Addition of 3D-EAM for navigation does not confer any benefits in terms of outcomes or procedure, with the exception of reduced irradiation, and is often associated with high conversion rates to fluoroscopy-based navigation. Further studies are necessary to establish non-inferiority of the PVI+PVA approach, potentially leading to a revision of the whole AF ablation framework.

## Data Availability

Data are available when required

## Non-standard Abbreviations and Acronyms

AF: Atrial fibrillation
PVs: pulmonary veins
PVI: Pulmonary vein isolation
PFA: Pulsed field ablation
3D-EAM: 3-dimensional electroanatomical mapping
PVA: Pulmonary vein ablation

## Acknowledgments

We would like to thank Dimitris Sougiannis, Stavros Koutsoubelis, Vasilios Michopoulos and Panagiotis Gkanas for their technical support

